# Race-Specific Dementia Risk Prediction Using 2024 Lancet Commission Risk Factors and Resting Heart Rate: A Survival Analysis of 55,004 NACC Participants

**DOI:** 10.64898/2026.08.23.26361129

**Authors:** Shakiru A. Alaka, So Fong Cam Ngan, Ranjith Iyappan, Jones Nwaeze, Bianca D’Amore, Maryam Katoueezadeh, Yazhini Thinakaran, Mahdi Hejazi Laein, Jake Baker, Siu Kwan Sze

## Abstract

Persistent racial disparities in dementia raise concerns regarding the validity and generalizability of existing prognostic models across diverse populations. We evaluated the utility of the 2024 Lancet Commission risk factors and resting heart rate (RHR) for race-specific dementia risk prediction using data from 55,004 participants in the National Alzheimer’s Coordinating Center cohort. Cox proportional hazards and Random Survival Forest (RSF) models were developed separately for Black, White, Asian, and American Indian participants to predict 1-, 3-, and 5-year time to dementia. RSF consistently outperformed Cox models across all racial groups and prediction horizons, achieving 5-year AUCs of 0.88-0.91 compared with 0.69-0.81 for Cox models. Inclusion of RHR modestly and consistently improved predictive performance across racial groups. Predictor importance varied between racial groups, suggesting heterogeneity in dementia risk profiles and disease presentation. These findings support the potential utility of RHR as a complementary prognostic biomarker and highlight the importance of equitable, personalized dementia risk prediction across diverse populations.

## 1 INTRODUCTION

Dementia imposes a substantial burden on patients, families, and health systems. Given the limited effective treatments, earlier identification of individuals at elevated risk is essential for implementing preventive strategies and reducing the disease burden.^1^ However, accurate dementia risk prediction remains a major clinical challenge due to its complex and heterogeneous nature.^2,3^ Therefore, new tools to support clinicians in decision making are critical for improving patient outcomes and delaying cognitive decline. In response, the medical community has identified several modifiable risk factors and developed risk prediction scores.^4,5^ These scores are typically rule based classifiers used in clinical practice.

However, persistent racial disparities in dementia raise important concerns regarding the fairness and generalizability of existing prognostic models, many of which have been developed using predominantly White populations.^6^ Although several racial and ethnic minority groups experience a disproportionately higher burden of dementia and cognitive impairment, these populations remain underrepresented in many longitudinal clinical and research cohorts.^6^ This imbalance may limit the ability of current prediction models to accurately estimate dementia risk across diverse populations and may contribute to unequal clinical decision-making and delayed intervention.^7^

The mechanisms underlying racial disparities in dementia are complex and multifactorial.^8^ Beyond genetic susceptibility, differences in educational opportunities, cardiovascular health, healthcare access, socioeconomic disadvantage, chronic stress exposure, and structural inequities are believed to substantially influence cognitive aging and dementia risk.^6–9^ Minority populations also frequently experience higher rates of vascular and cardiometabolic conditions, including hypertension, diabetes, obesity, and cardiovascular disease, all of which are established contributors to cognitive decline.^10–12^ These factors are often inadequately captured or incompletely modeled in existing prognostic frameworks.^13^

Age remains the strongest known risk factor for dementia, yet the interaction between aging, race, and modifiable risk factors is not fully understood.^14^ Most prognostic models are developed in older cohorts where age-related effects may dominate other clinically relevant predictors, potentially masking subgroup-specific risk patterns.^15^ In addition, differences in follow-up duration, healthcare utilization, and diagnostic pathways, across racial groups may further affect the accuracy and calibration of dementia prediction models over time.^14–16^

Current dementia prognostic studies have also largely focused on traditional statistical approaches and aggregate measures of predictive performance, with limited attention to subgroup fairness and calibration across racial populations.^17^ While Cox proportional hazards regression remains widely used because of its interpretability, machine learning survival methods such as random survival forests may better capture complex non-linear relationships and interactions among demographic, cardiovascular, and clinical variables.^17^ These approaches may be particularly valuable in heterogeneous populations where risk patterns are unlikely to follow simple linear assumptions.

Recent public health frameworks, including the 2024 Lancet Commission, have emphasized the importance of modifiable risk factors in dementia prevention and progression.^18,19^ Incorporating these factors alongside physiological markers such as resting heart rate may improve individualized risk prediction and provide additional insight into the relationship between cardiovascular health and cognitive decline.^20,21^ However, few studies have systematically evaluated the performance of survival-based models for early dementia risk prediction and risk stratification across racial groups over clinically relevant timeframes.^22,23^

This study aimed to develop and compare Cox proportional hazards and random survival forest models for predicting 1-, 3-, and 5-year dementia risk based on Lancet Commission risk domains and resting heart rate using data from the U.S. National Alzheimer’s Coordinating Center cohort. We further evaluated model performance across racial groups to assess the robustness, predictive accuracy, and potential equity of these approaches in diverse aging populations. We hypothesized that the predictive utility and relative importance of dementia risk factors would differ across racial groups and that inclusion of resting heart rate would improve risk prediction beyond established Lancet Commission risk factors.

## 2 METHODS

We conducted a prognostic modeling study using prospectively collected longitudinal cohort data from the National Alzheimer’s Coordinating Center (NACC) Uniform Data Set version 3.0. ^24^ The study is reported in line with the “transparent reporting of a multivariable prediction model for individual prognosis or diagnosis (TRIPOD) principle.^25,26^ The completed checklists is provided in supplementary Table S1.

### 2.1 Data Source

We utilized data from the NACC Uniform Data Set (UDS) version 3.0,^27^ which includes participant visits collected between September 1, 2005, and December 31, 2025. The dataset comprised 205,908 participant visits. The data were collected at 39 Alzheimer’s Disease and Research Centers (ADRC), predominantly in metropolitan regions across the United States. Individuals were recruited through clinical referrals or through active community recruitment process that comply with the ADRC guidelines.^24^ The dataset includes individuals’ demographic, clinical characteristics, family history, medical history, types of medication use, and neurophysiological assessment individuals underwent during annual follow-up visits with comprehensive clinical assessments until death or drop out.^24,28^

Each Alzheimer’s Disease Center (ADC) contributing to the National Alzheimer’s Coordinating Center obtains informed consent from participants and approval from its local Institutional Review Board (IRB) before submitting data.^24^ For this study, access to the dataset was granted through an approved data request to NACC. In addition, this study received ethics approval from the Brock University Health Sciences Research Ethics Board (REB # 24072). All data used were de-identified prior to analysis, and details of the dataset have been described elsewhere.

### 2.2 Survival Analysis Dataset

The dataset was developed by transforming the longitudinal NACC visit data into a person-level format suitable for time-to-event analysis. Records were first sorted by participant ID and follow-up time (naccfdays) to ensure proper temporal ordering. For each unique ID, the time variable was defined as the earliest recorded time of dementia diagnosis if it occurred, or the last available follow-up time otherwise. An event indicator was then created, coded as 1 for incident dementia and 0 for censored observations. The dataset was subsequently reduced to one row per participant, preserving each individual’s follow-up trajectory in terms of time and event status. Participant with dementia at baseline were excluded, and death before dementia was treated as a censoring event.

### 2.3 Data Preprocessing

To ensure data quality and consistency, all special missing-value codes were recoded as missing (NA). Specifically, values indicating “Not Available” (−4 and 4.4), “Not accessed” or “Not applicable” (8, 88, 888.8, and 8888), and “Unknown” (9, 99, 999, 999.9, and 9999) were converted to NA according to variable-specific coding schemes. In addition, blank or empty entries across all variables were also recoded as NA. Variables with inconsistent or non-standard coding (for example, RACE, where code 50 represents “Other”) were carefully reviewed and manually recoded to NA to ensure consistency. Finally, all categorical variables were converted from numeric formats to factor variables to facilitate appropriate statistical handling.

### 2.4 Assessment of Lancet Commission Risk Factors and Resting Heart Rate

For this study, variables required to estimate the time to dementia were collected during standardized clinical assessments at Alzheimer’s Disease Research Centers (ADRCs) contributing to the National Alzheimer’s Coordinating Center (NACC) database.^24^ Predictors were selected to reflect Lancet Commission risk domains available in NACC. Age (years) and sex (male/female) were documented for each participant at the time of the visit.^29^ Educational attainment (years of formal schooling) was self-reported by participants or their informants using structured interviews.^29^ Systolic blood pressure (SBP, mmHg) was measured by trained staff using calibrated sphygmomanometers while participants were seated and rested; multiple measurements were averaged to ensure accuracy. In addition, a resting heart rate (RHR, beats per minute) was recorded during the same visit, either by pulse palpation or via electrocardiogram after the participant had rested. Body mass index (BMI, kg/m²) was calculated from standardized weight and height measurements taken by trained personnel with participants in light clothing and without shoes.^29^ Total cholesterol (mg/dL) was obtained from fasting blood samples and analyzed according to standard laboratory procedures Traumatic brain injury, hearing loss, and diabetes mellitus were ascertained through clinician diagnosis, medical records, and participant or informant report using structured study interviews. Where available, objective clinical data were used to support case definitions, including fasting plasma glucose (mg/dL) or glycated haemoglobin (HbA1c, %) for diabetes mellitus. Hearing loss and traumatic brain injury were identified based on documented clinical history, as no standardised laboratory units apply to these conditions.^29^ The variables used in this study and their corresponding descriptors are available in (Supplementary Table S2).

### 2.5 Statistical Analysis

Descriptive statistics (median, and interquartile range, frequencies, and percentages) were used to summarize individual demographic and clinical characteristics by time to dementia status for four different race datasets. Univariate associations between time to dementia status and each categorical and continuous predictor variable were assessed using the chi-square test and Wilcoxon rank-sum tests, respectively. The predictors included in the analyses exhibited substantial missingness, ranging from 10% to 60% across both categorical and continuous variables (Supplementary Figure S1). To address this, multivariate imputation by chained equation (MICE)^30,31^ was applied to reduce the impact of missing data and minimize potential bias. In addition, given the slight class imbalance (fewer individuals developing dementia than remaining dementia-free), oversampling of the minority class was applied to mitigate the potential impact of class imbalance on model performance.

Figure S2 presents the workflow schematic of the whole process. Time to dementia risk prediction models were developed separately for each racial group using random forest survival^32^ and Cox proportional hazard models.^33^ Each model was developed using patients’ demographic and clinical characteristics, including age, education, hypertension, hearing loss, body mass index, cholesterol, obesity, depression, diabetes, traumatic brain injury, and resting heart rate, as input variables to predict time to dementia among individual with cognitive impairment. For each of the racial datasets, the difference the in-survival distributions were accessed using log-rank tests. Multivariable Cox proportional hazards regression models were fitted to examine association and estimate the hazard ratios (HRs) and 95% confidence intervals (CIs) between risk factors and dementia by race. The proportional hazards assumption was evaluated using Schoenfeld residuals method.

Random Survival Forest (RSF) models were additionally implemented within each racial dataset to account for potential nonlinear associations and higher-order interactions among predictors. In addition, hyper parameter tuning process was used for these models. For example, the number of trees in random survival forest was adjusted from 500 to 5000 and the number of variables for splitting at each tree node was at the interval between 3 to 5. This process is to eliminate the chance of overfitting and underfitting and compare its performance in order to determine which set of hyper parameters results in the most accurate.^32^ Model performance was assessed using Harrell’s concordance index (C-index),^34^ integrated brier score (IBS),^35^ and time-dependent predictive performance at 1-, 3-, and 5-year follow-up intervals. While higher values on AUC indicate better predictive accuracy, higher values of Integrated Brier score (>0.25) indicate poor discrimination/calibration of the model. Furthermore, variable importance measures were used to identify the strongest predictors of dementia risk within each racial group. These were achieved using the R software’s *fastshap* function in *SHAP* package.^36^ All the analyses were conducted in R software.^37^ Statistical significance was evaluated at alpha = 0.05. A sample of this code is presented as a supplementary with the manuscript.

## 3 RESULTS

### 3.1 Cohort Characteristics and Dementia Risk Factors

Table 1 presents the complete-case demographic and clinical characteristics of participants across the Black, White, Asian, and American Indian cohorts. There were significant differences in demographic and clinical profiles among the four groups. Table 2 presents the demographic and clinical characteristics of individuals with and without dementia in the NACC study. Of the 55,004 participants, 17,403 (46.3%) were classified as having dementia. Individuals with dementia were more likely to be older (p < 0.01), female (p < 0.01), and to have higher resting heart rates. They were also more likely to report current antidepressant use (p < 0.01), and a history of traumatic brain injury (p < 0.01) compared with individuals without dementia. Figure 1 describes the association between the risk factors and dementia by race. Increasing age and diabetes, particularly type I diabetes, were among the strongest predictors of dementia across racial groups, except in the Black cohort. Depression also showed consistent positive associations with dementia across all racial groups.

**Figure 1.**
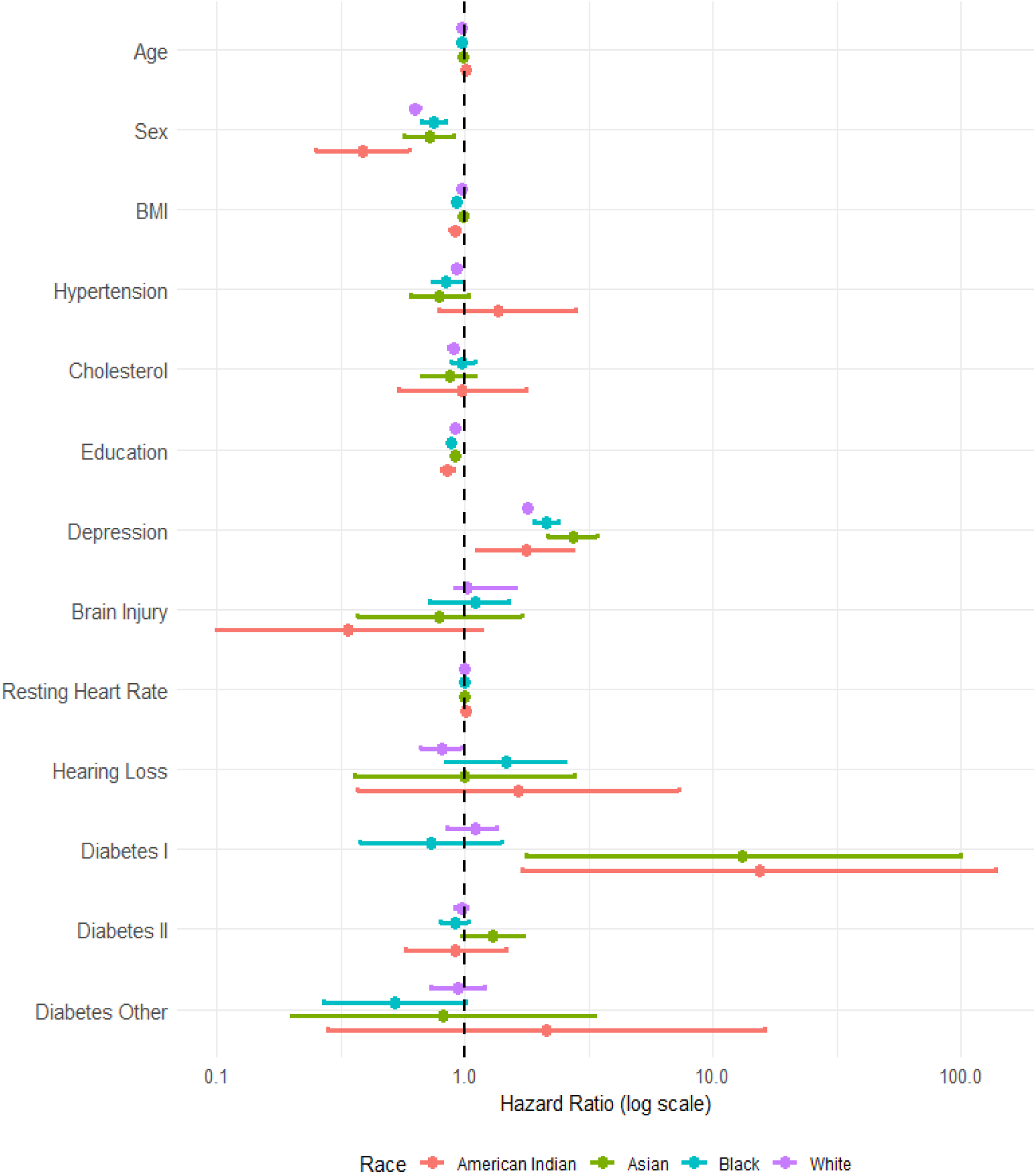
Racial variation in the association between dementia risk factors and incident dementia. Hazard ratios and 95% confidence intervals for demographic and clinical predictors included in the Lancet Commission framework are shown for White, Black, Asian, and American Indian participants. Although age remained the most consistent predictor across all groups, the magnitude of associations for several risk factors varied by race. BMI = body mass index; HR = hazard ratio; NACC = National Alzheimer’s Coordinating Center.

**Table 1.** Baseline demographic and clinical characteristics of participants by racial group in the NACC cohort.

| Participant Characteristics | White<br>N = 43,030 | Black<br>N = 8,484 | Asian<br>N = 1,543 | American Indian<br>N = 533 | P - value |
| --- | --- | --- | --- | --- | --- |
| Age, median (IQR) | 76 (68, 82) | 74 (67, 81) | 73 (66, 80) | 69 (63, 77) | <0.001 |
| Sex |  |  |  |  | <0.001 |
| Male (n, %) | 19,744 (46%) | 2,364 (28%) | 625 (41%) | 175 (33%) |  |
| Female (n, %) | 23,286 (54%) | 6,120 (72%) | 918 (59%) | 358 (67%) |  |
| Education, median (IQR) | 16 (13, 18) | 14 (12, 16) | 16 (15, 18) | 12 (12, 15) | <0.001 |
| Hypertension |  |  |  |  | <0.001 |
| No (n, %) | 22,772 (53%) | 2,298 (27%) | 814 (53%) | 227 (43%) |  |
| Yes (n, %) | 20,258 (47%) | 6,186 (73%) | 729 (47%) | 306 (57%) |  |
| BMI, median (IQR) | 25.9 (23.2, 29.3) | 28.2 (24.7, 32.6) | 23.4 (21.2, 26.1) | 28.5 (25.3, 34.5) | <0.001 |
| Cholesterol |  |  |  |  | <0.001 |
| No (n, %) | 19,709 (46%) | 3,361 (40%) | 671 (43%) | 203 (38%) |  |
| Yes (n, %) | 23,321 (54%) | 5,123 (60%) | 872 (57%) | 330 (62%) |  |
| Resting Heart Rate, median (IQR) | 68 (60, 76) | 69 (61, 76) | 69 (62, 76) | 68 (61, 76) |  |
| Depression |  |  |  |  | <0.001 |
| No (n, %) | 26,717 (62%) | 6,845 (81%) | 1,249 (81%) | 383 (72%) |  |
| Yes (n, %) | 16,313 (38%) | 1,639 (19%) | 294 (19%) | 150 (28%) |  |
| Brain Injury |  |  |  |  | <0.001 |
| No (n, %) | 42,086 (98%) | 8,319 (98%) | 1,517 (98%) | 507 (95%) |  |
| Yes (n, %) | 944 (2.2%) | 165 (1.9%) | 26 (1.7%) | 26 (4.9%) |  |
| Hearing Loss |  |  |  |  | 0.007 |
| No (n, %) | 42,707 (99%) | 8,437 (99%) | 1,527 (99%) | 524 (98%) |  |
| Yes (n, %) | 323 (0.8%) | 47 (0.6%) | 16 (1.0%) | 9 (1.7%) |  |
| Diabetes |  |  |  |  | <0.001 |
| No Diabetes (n, %) | 37,765 (88%) | 6,040 (71%) | 1,228 (80%) | 358 (67%) |  |
| Type I (n, %) | 265 (0.6%) | 72 (0.8%) | 3 (0.2%) | 3 (0.6%) |  |
| Type II (n, %) | 4,793 (11%) | 2,312 (27%) | 294 (19%) | 171 (32%) |  |
| Other Type (n, %) | 207 (0.5%) | 60 (0.7%) | 18 (1.2%) | 1 (0.2%) |  |

Values are presented as median interquartile range (IQR) or n (%). P values were calculated using Kruskal-Wallis tests for continuous variables and $\chi^2$ tests for categorical variables; BMI (Body Mass Index). NACC = National Alzheimer's Coordinating Center.

**Table 2.** Baseline characteristics of participants with and without incident dementia.

| Participant Characteristics | No Dementia<br>(N = 37, 601) | Dementia<br>(N = 17,403) | P - value |
| --- | --- | --- | --- |
| Age, median (IQR) | 71 (65, 77) | 73 (65, 80) | <0.001 |
| Sex |  |  | <0.001 |
| Male (n, %) | 15,006 (40%) | 8,393 (48%) |  |
| Female (n, %) | 22,595 (60%) | 9,010 (52%) |  |
| Education |  |  | <0.001 |
| < Secondary (n, %) | 6,858 (18%) | 5,272 (30%) |  |
| High School (n, %) | 582 (1.5%) | 663 (3.8%) |  |
| Post Secondary (n, %) | 30,161 (80%) | 11,468 (66%) |  |
| Hypertension |  |  | 0.900 |
| No (n, %) | 19,430 (52%) | 8,991 (52%) |  |
| Yes (n, %) | 18,171 (48%) | 8,412 (48%) |  |
| Body Mass Index |  |  | <0.001 |
| <30 (n, %) | 27,159 (72%) | 13,803 (79%) |  |
| >30 (n, %) | 10,442 (28%) | 3,600 (21%) |  |
| Cholesterol |  |  | 0.800 |
| No (n, %) | 17,731 (47%) | 8,224 (47%) |  |
| Yes (n, %) | 19,870 (53%) | 9,179 (53%) |  |
| Resting Heart Rate |  |  | <0.001 |
| Normal (60–100 bpm) | 30,407 (81%) | 14,091 (81%) |  |
| Low (<60 bpm) | 6,954 (18%) | 3,141 (18%) |  |
| Higher (>100 bpm) | 240 (0.6%) | 171 (1.0%) |  |
| Race |  |  | <0.001 |
| White (n, %) | 29,102 (77%) | 14,929 (86%) |  |
| Black (n, %) | 6,861 (18%) | 1,907 (11%) |  |
| American Indian (n, %) | 395 (1.1%) | 156 (0.9%) |  |
| Native Hawaiian (n, %) | 33 (<0.1%) | 31 (0.2%) |  |
| Asian (n, %) | 1,210 (3.2%) | 380 (2.2%) |  |
| Depression |  |  | <0.001 |
| No (n, %) | 28,548 (76%) | 10,113 (58%) |  |
| Yes (n, %) | 9,053 (24%) | 7,290 (42%) |  |
| Brain Injury |  |  | 0.300 |
| No (n, %) | 36,893 (98%) | 17,052 (98%) |  |
| Yes (n, %) | 708 (1.9%) | 351 (2.0%) |  |
| Hearing Loss |  |  | 0.600 |
| No (n, %) | 37,419 (100%) | 17,325 (100%) |  |
| Yes (n, %) | 182 (0.5%) | 78 (0.4%) |  |
| Diabetes |  |  | 0.200 |
| No Diabetes (n, %) | 31,769 (84%) | 14,797 (85%) |  |
| Type I (n, %) | 262 (0.7%) | 107 (0.6%) |  |
| Type II (n, %) | 5,314 (14%) | 2,397 (14%) |  |
| Other Type (n, %) | 256 (0.7%) | 102 (0.6%) |  |

IQR = interquartile range; Bpm = beats per minutes; N = Sample size

### 3.2 Predictive Performance of Survival Models Across Racial Groups

Table 3 describes the predictive accuracy of random survival forest and Cox proportional hazard regression algorithms for predicting 1-,3-and 5-year time to dementia across Black, White, Asian and American Indian racial groups using the Lancet features with resting heart rate. There were significant differences in the predictive accuracy of between both models, regardless of the racial groups. For example, for models trained using the Lancet features with resting heart rate, RSF achieved excellent discrimination across all racial groups, including populations that are often underrepresented in dementia prediction studies, suggesting that machine-learning survival models may better accommodate heterogeneous risk structures than traditional regression approaches. Specifically, the performance improves at longer time horizons (especially at 5-year) with average range of AUC 0.88 (95%CI = [0.86, 0.91]) and 0.91 (95%CI = [0.86, 0.95]) while 1-year predictive discrimination had the least average AUC range of 0.80 (95%CI = [0.71,0.87]) and 0.83 (95%CI = [0.79, 0.86]) across groups (See figure 2). Table 3 describes the predictive accuracy for the models trained using Cox proportional hazard model and same set of features, the model shows a moderate discriminative performance. Specifically, the performance was slightly with longer follow up but remain below random forest survival at 5-year AUC range of 0.69 (95%CI = [0.64, 0.73]) and 0.81(95%CI = [0.75, 0.87]) while 1-year predictive discrimination had the least AUC range of; 0.64 (95%CI = [0.61, 0.67]) and 0.75 (95%CI = [0.68, 0.83]) across racial groups (See figure 3).

**Figure 2.**
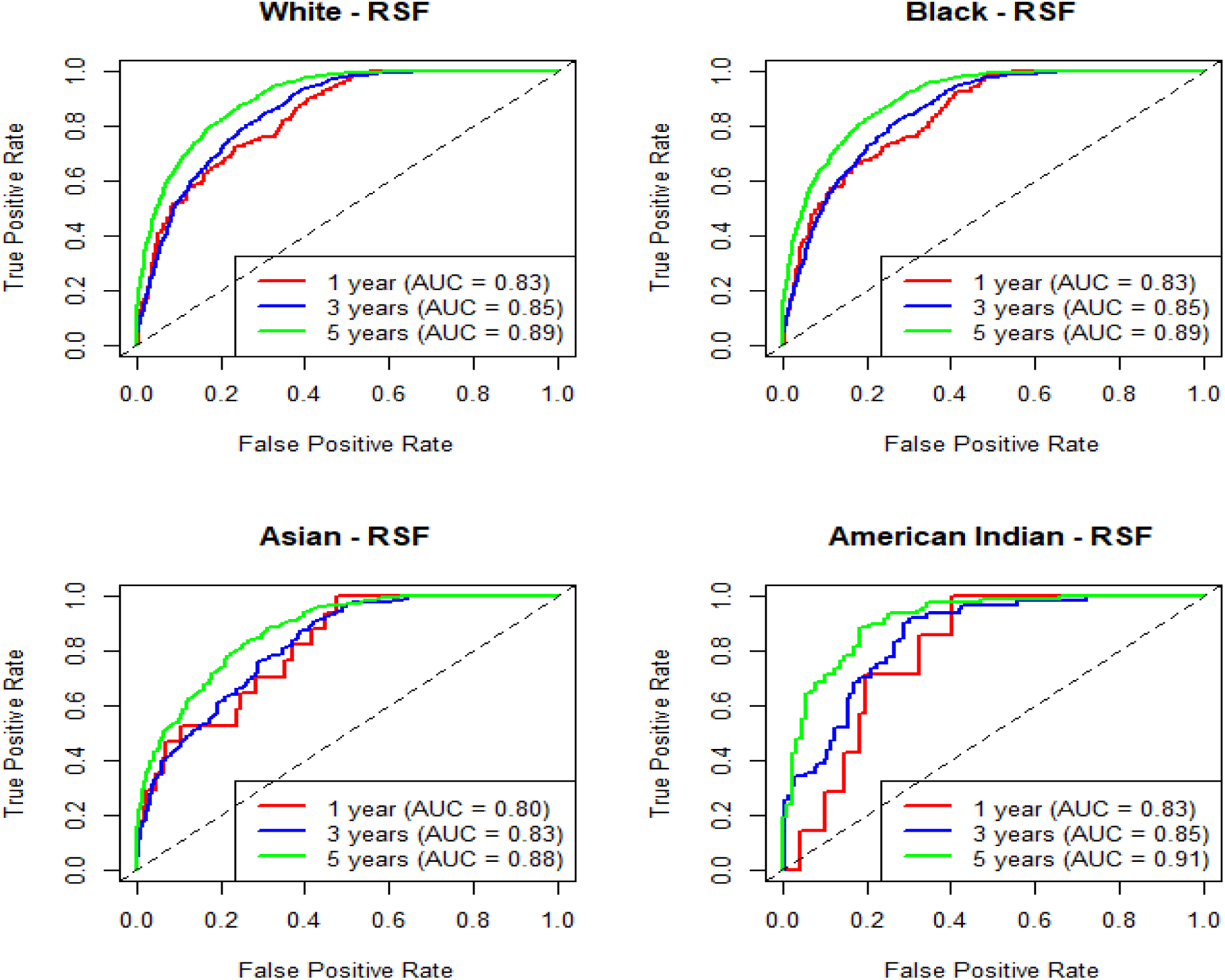
Random Survival Forest Models Demonstrate Strong Dementia Risk Prediction Across Racial Groups. Time-dependent receiver operating characteristic (ROC) curves for Random Survival Forest (RSF) models predicting 1-, 3-, and 5-year dementia risk using Lancet Commission risk factors and resting heart rate. RSF models achieved strong discriminative performance across all racial groups, with predictive accuracy generally improving at longer follow-up periods. Five-year predictions consistently showed the highest area under the curve (AUC) values, indicating that long-term dementia risk can be estimated with high accuracy using readily available clinical variables. AUC = area under the receiver operating characteristic curve; NACC = National Alzheimer’s Coordinating Center; ROC = receiver operating characteristic; RSF = Random Survival Forest.

**Figure 3.**
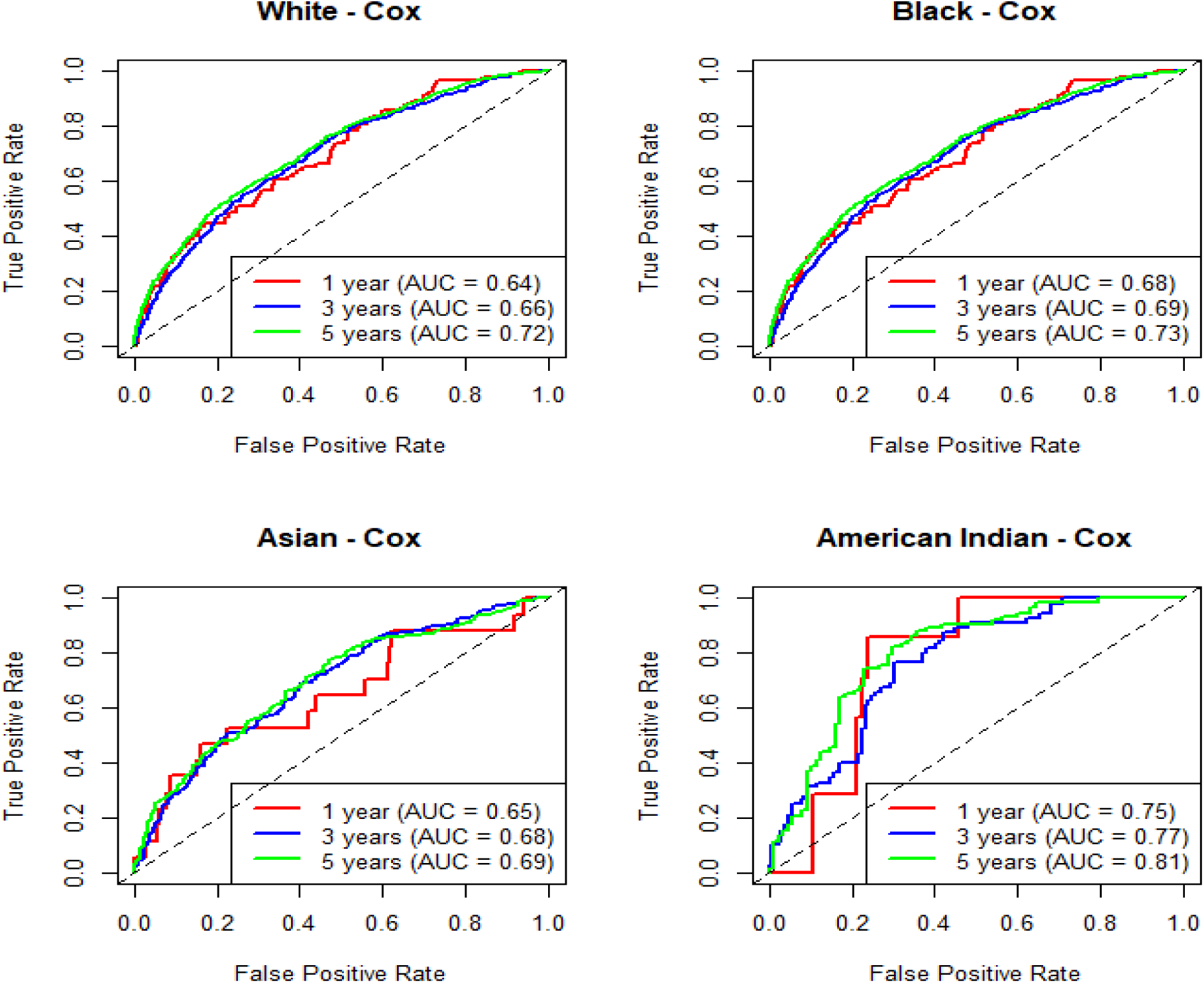
Cox Proportional Hazards Models Predict Dementia Risk Across Racial Groups. Time-dependent receiver operating characteristic (ROC) curves for Cox proportional hazards models predicting 1-, 3-, and 5-year dementia risk using Lancet Commission risk factors and resting heart rate. Cox models demonstrated moderate predictive performance across racial groups, with discrimination improving at longer follow-up periods. Although predictive accuracy was lower than that observed for Random Survival Forest models, the results indicate that conventional regression-based approaches remain useful for estimating dementia risk in diverse populations. AUC = area under the receiver operating characteristic curve; NACC = National Alzheimer’s Coordinating Center; ROC = receiver operating characteristic.

**Table 3.** Time-dependent predictive performance of Random Survival Forest and Cox proportional hazards models using Lancet Commission risk factors and resting heart rate across racial groups.

| Random Survival Forest |  |  |  |  |
| --- | --- | --- | --- | --- |
| Survival Threshold | White | Black | Asian | American Indian |
| One-year | 0.83 (0.79, 0.86) | 0.83 (0.79, 0.86) | 0.80 (0.71, 0.87) | 0.83 (0.74, 0.92) |
| Three-year | 0.85 (0.84, 0.86) | 0.85 (0.84, 0.86) | 0.83 (0.79, 0.86) | 0.85 (0.80, 0.91) |
| Five -year | 0.89 (0.88, 0.91) | 0.89 (0.88, 0.91) | 0.88 (0.86, 0.91) | 0.91 (0.86, 0.95) |
| Cox Proportional Hazard |  |  |  |  |
| One-year | 0.64 (0.61, 0.67) | 0.68 (0.63, 0.74) | 0.68 (0.54, 0.81) | 0.75 (0.68, 0.83) |
| Three-year | 0.66 (0.63, 0.68) | 0.69 (0.68, 0.72) | 0.66 (0.61, 0.71) | 0.77 (0.71, 0.84) |
| Five -year | 0.72 (0.69, 0.75) | 0.73 (0.71, 0.75) | 0.69 (0.64, 0.73) | 0.81 (0.75, 0.87) |

Values are presented as AUC (95% confidence interval). AUC = area under the receiver operating characteristic curve; CI = confidence interval.

### 3.3 Impact of Resting Heart Rate on Model Performance

When the models were refit without resting heart rate, their predictive performance was generally slightly reduced or remained comparable to the models that included resting heart rate (Figure 4 & table 4). However, these differences in predictive accuracy were statistically significant across all racial groups. For example, at the 5-year time point, the random survival forest maintained strong discrimination even without resting heart rate, with AUCs of 0.88 (95%CI = [0.87, 0.89]), 0.88 (95%CI = [0.87, 0.89]), 0.86 (95%CI = [0.83, 0.89]), and 0.88 (95%CI = [0.84, 0.94]) for Black, White, Asian, and American Indian cohorts, respectively. In contrast, the Cox proportional hazards model showed lower performance, with AUCs ranging approximately from 0.68 (95%CI = [0.64, 0.73]), to 0.79 (95%CI = [0.75, 0.87]) when resting heart rate was excluded from the predictor set. (See figure 5 & table 4). Although the absolute improvements in discrimination were modest, the consistent performance gains observed across racial groups and modeling approaches suggest that resting heart rate provides complementary prognostic information beyond traditional dementia risk factors. On the other hand, the rank ordering of predictors according to their relative contribution to the prediction of time to dementia is largely dependent on the racial groups (Figure 6). SHAP analysis demonstrated substantial variation in predictor importance across racial groups. Although age was consistently the most influential predictor, the relative contributions of education, depression, diabetes, cholesterol, hypertension, body mass index, and resting heart rate varied considerably between groups.

**Figure 4.**
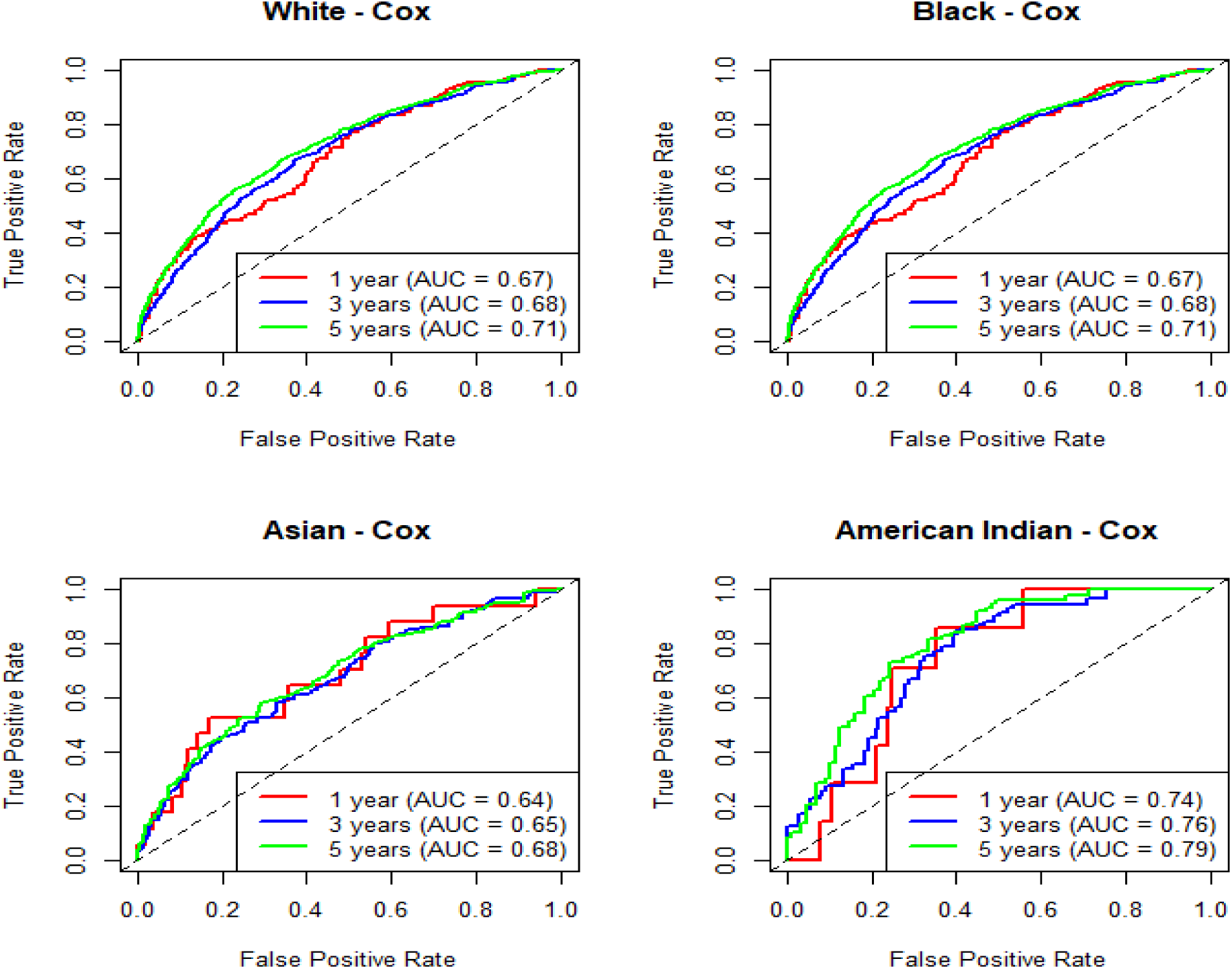
Impact of Excluding Resting Heart Rate on Cox Model Performance. Time-dependent receiver operating characteristic (ROC) curves for Cox proportional hazards models predicting 1-, 3-, and 5-year dementia risk using Lancet Commission risk factors without resting heart rate. Exclusion of resting heart rate resulted in a modest reduction in predictive performance across most racial groups compared with models that included resting heart rate. Despite this reduction, model discrimination generally improved with increasing follow-up duration, with the highest AUC values observed at 5 years. AUC = area under the receiver operating characteristic curve; NACC = National Alzheimer’s Coordinating Center; ROC = receiver operating characteristic.

**Figure 5.**
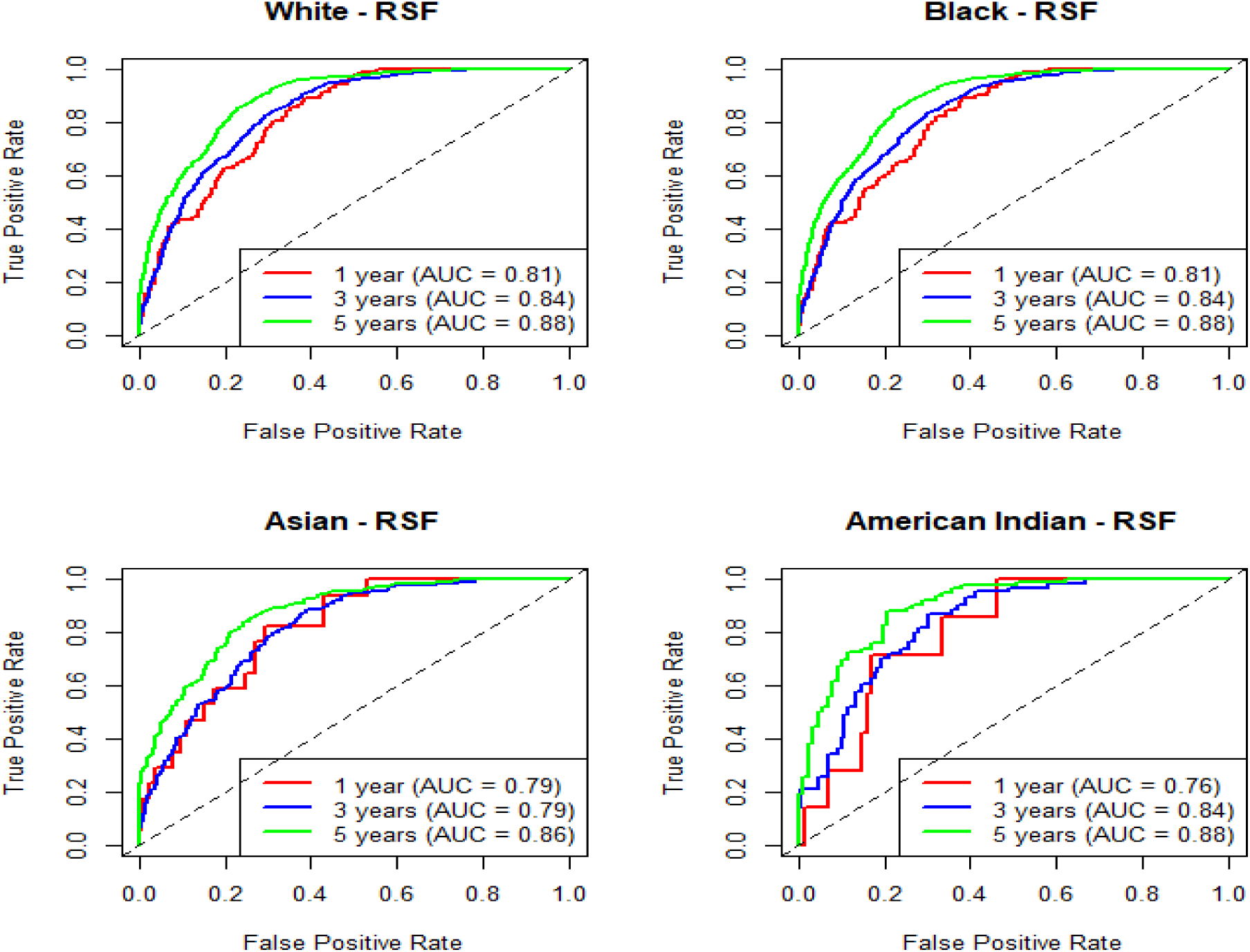
Random Survival Forest Models Maintain Strong Performance Without Resting Heart Rate. Time-dependent receiver operating characteristic (ROC) curves for Random Survival Forest models predicting 1-, 3-, and 5-year dementia risk using Lancet Commission risk factors without resting heart rate. Removal of resting heart rate resulted in only a modest reduction in predictive accuracy, indicating that RSF models remained robust across racial groups. Predictive discrimination remained highest at the 5-year follow-up interval, supporting the utility of machine learning approaches for long-term dementia risk prediction. AUC = area under the receiver operating characteristic curve; NACC = National Alzheimer’s Coordinating Center; ROC = receiver operating characteristic; RSF = Random Survival Forest.

**Figure 6.**
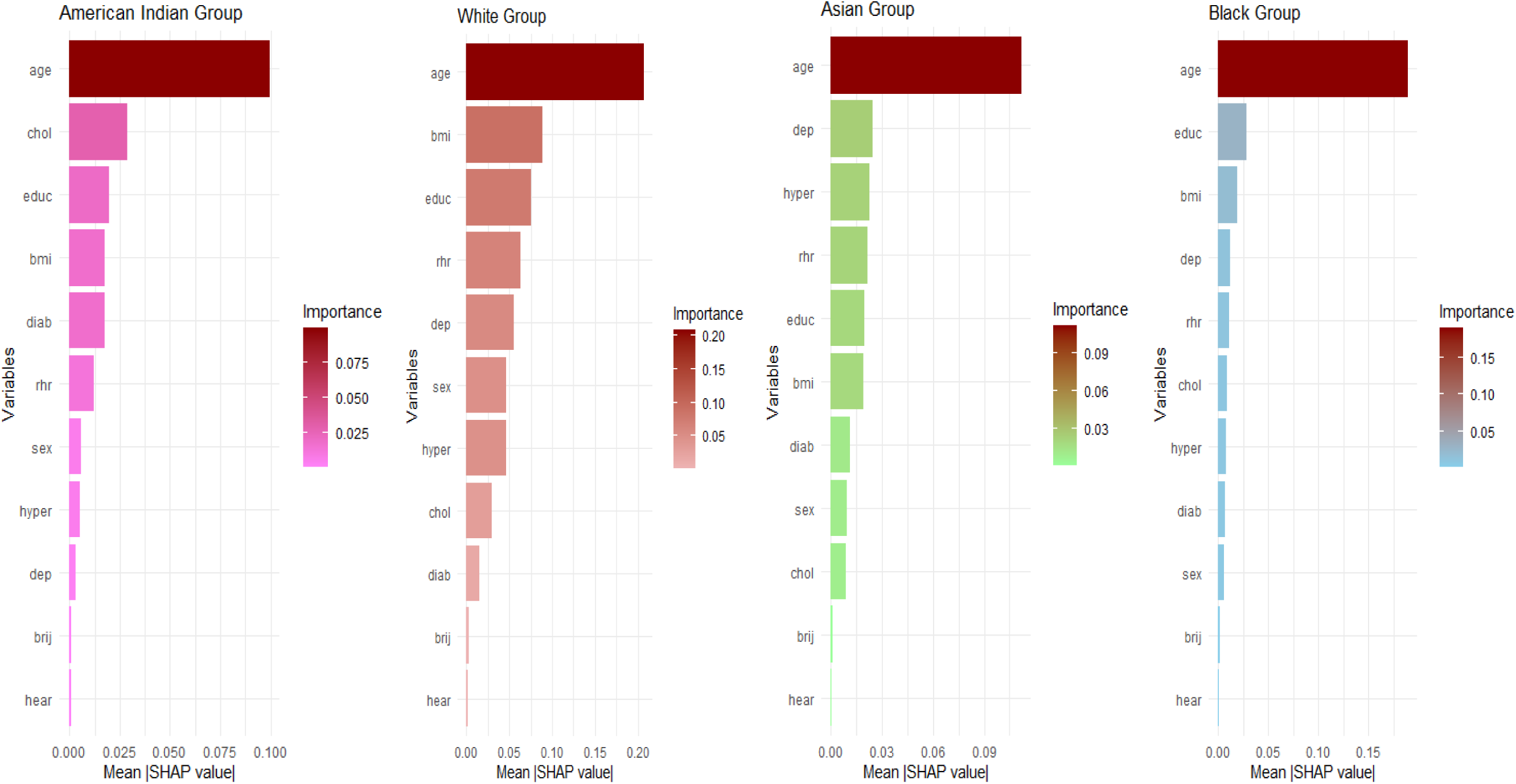
Relative importance of dementia risk factors across racial groups determined using SHAP analysis. Mean absolute SHAP (SHapley Additive exPlanations) values derived from Random Survival Forest models were used to quantify the contribution of each predictor to dementia risk prediction. Age was consistently the most influential predictor across all racial groups. However, the relative importance of education, depression, diabetes, cholesterol, hypertension, body mass index, and resting heart rate differed substantially between racial groups, suggesting heterogeneity in dementia risk profiles and potentially distinct pathways contributing to dementia risk.

**Table 4.**
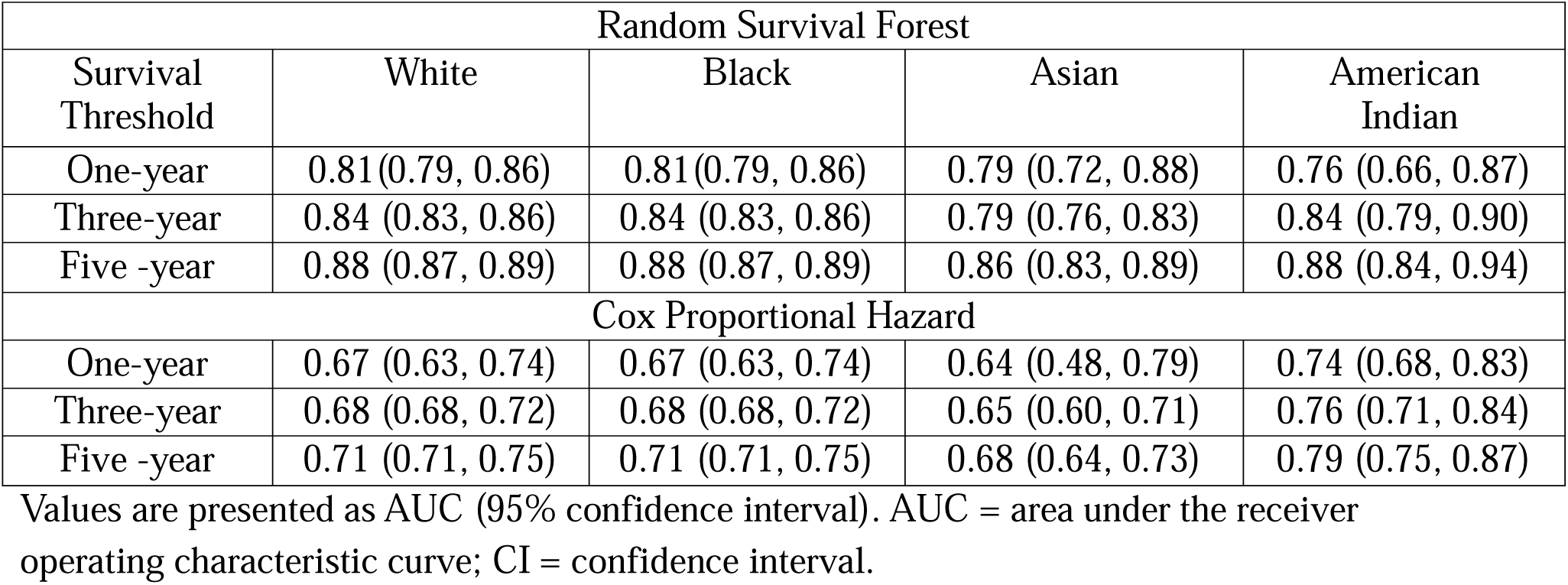
Time-dependent AUCs of Random Survival Forest and Cox proportional hazards models using Lancet Commission risk factors without resting heart rate across racial groups.

### 3.4 Model Calibration and Integrated Brier Score Analysis

Integrated brier scores (IBS) were used to assess the calibration performance of all the investigated in the Black, White, Asian and American Indian cohorts, respectively. There were no significant differences in IBS scores for random survival forest and regression-based models across all cohorts, regardless of the including resting heart rate variable or without (Figure S3, table S3 & S4). A perfectly performing model would have an Integrated Brier Score (IBS) of 0, indicating no prediction error across all time points. As the IBS increases, the model’s predictive accuracy worsens, reflecting greater deviation between predicted probabilities and observed outcomes over time. Thus, the higher the IBS, the poorer the overall model performance, while lower values indicate better calibration and discrimination. The generally low IBS values observed across racial groups indicate low overall prediction error and suggest good agreement between predicted and observed dementia outcomes across populations and prediction horizons. For example, when models were developed with resting heart rate, RSF exhibited relatively low and stable IBS values across racial groups, with only modest increases from 1-year to 5-year predictions. In contrast, the Cox model showed higher IBS values and greater variability across groups, indicating comparatively poorer performance. Notably, some racial groups (e.g., Asian and American Indian cohorts) showed slightly higher IBS at longer time horizons, suggesting reduced accuracy in these subgroups. When resting heart rate was excluded, both models showed a slight increase in IBS, indicating a modest decline in predictive performance. However, this deterioration was more pronounced for the Cox model, while RSF remained relatively robust, with only minimal changes in IBS across time points and racial groups.

### 3.5 Predictor Importance and Dementia Risk Stratification

We evaluated the clinical utility of the models by examining the derived dementia risk scores. The aim was to determine whether each model meaningfully stratified participants into clinically interpretable risk groups, rather than focusing solely on overall discrimination and calibration. Participants were categorized into three clinical risk groups (low, medium, and high) based on their predicted scores. For example, higher risk scores were increasingly concentrated in the high-risk group (see Figure S4, left panel). Furthermore, individuals with higher risk scores tended to develop dementia earlier, whereas those with lower risk scores were more likely to remain dementia-free over the follow-up period (see Figure S4, right panel). Overall, the distribution of risk scores was concentrated in the mid-range (30–60) and appeared approximately symmetric, suggesting no substantial skewness (See figure S5).

## 4 DISCUSSIONS

This study has three main findings. First, Random Survival Forest models consistently outperformed Cox proportional hazards models across all racial groups. Second, resting heart rate provided modest but consistent improvements in predictive performance. Third, the relative importance of dementia risk factors differed across racial groups, suggesting heterogeneity in dementia risk profiles.

This study developed and compared machine learning model and regression-based model for predicting time to dementia using 55,004 unique NACC individuals in the United States from 2005 to 2025. Our analyses revealed that random survival forest have a significant utility in the prediction of time to dementia across racial groups. First, we examined the association between the Lancet risk factors and dementia by race. Subsequently, we developed Lancet feature models including resting heart rate to predict time to dementia across four racial groups (Black, White, Asian and American Indian) at 3 distinct survival time-thresholds, all of which achieved high predictive performance. We additionally ranked predictors according to their relative contribution to prediction across the four racial groups and found that age remained the most important predictor in all groups. These time-to-dementia predictors highlight both similarities and differences in the underlying etiology and clinical presentation among individuals from different racial backgrounds.

The results of our analysis examining the association between Lancet risk factors and dementia across racial groups showed that increasing age,^15^ diabetes (type I),^38^ and depression^39^ were consistently associated with a higher risk of dementia. These findings are in line with previously established modifiable and non-modifiable risk factors reported in the literature.^16,17,40^ With respect to racial differences, relatively few studies have investigated the association between Lancet risk factors and dementia across diverse racial groups.^23,41,42^ Among the studies identified, key distinctions were observed in cohort composition and time-to-dementia outcomes across Black, White, and Asian populations. Our findings further support the presence of differences in underlying etiology and clinical presentation among individuals from different racial backgrounds.^6,8,10,43^

Subsequently, leveraging this insight, we developed random survival forest and Cox proportional hazards models using established Lancet risk factors, with and without inclusion of resting heart rate, to predict time to dementia at 1-, 3-, and 5-year horizons across four racial groups. The models demonstrated strong discriminative performance, with AUC values exceeding 0.88 at the 5-year time point across all racial groups. Performance was reduced at the 1-year timeframe, suggesting improved stability and predictive reliability for longer-term dementia risk estimation. Comparable patterns were observed for Cox proportional hazards models. Importantly, resting heart rate emerged as a previously underutilized biomarker that consistently improved predictive performance across both random survival forest and Cox proportional hazards models.^20,21^ This suggests that resting heart rate may capture multidimensional aspects of dementia risk, including cardiovascular integrity, autonomic nervous system function, and systemic physiological stress.^44^ Elevated resting heart rate has been linked to vascular dysfunction, reduced cerebral perfusion, and arterial stiffness, all of which are established contributors to cognitive decline and dementia pathophysiology.^45^

To understand, interpret, and gain trust in the random survival forest technique, we ranked the essential predictors with the highest contribution to the accuracy of these predictions by ranking them within each racial dataset (Figure 6). These predictors were explored to make certain they are in line with clinical domain knowledge, reasonable expectations, and offer insight into the importance of individual predictors for prediction performance. These variables importance takes the interaction between variables into account. Our evaluation of predictor importance across random survival forest demonstrated that age was the most influential predictor in predicting time-to-dementia outcomes across all racial groups. This finding was consistent across both modeling approaches and aligns with existing prognostic risk scores for dementia, which consistently identify age as the strongest predictor of dementia-related outcomes.^14,15^ Hence, the random survival forest could be used as an efficient screening tool to pick up predictive patterns in the data that could potentially lead to further hypothesis-driven research.

To our knowledge, this study is among the first to evaluate machine learning-based survival models for predicting time to dementia by incorporating resting heart rate with the Lancet commission risk factors across four racial groups. The findings suggest that resting heart rate may provide additional prognostic information to established dementia risk factors and also demonstrated the potential of machine learning approaches to support more personalized dementia risk assessment. The strong predictive performance observed at the 5-year horizon suggests that dementia risk can be identified well in advance, providing a meaningful window for early intervention and prevention strategies. Incorporating readily available measures such as resting heart rate into routine clinical risk models may enhance early risk stratification without substantially increasing clinical burden, particularly in primary care settings where scalable tools are essential. Beyond its predictive utility, resting heart rate may capture aspects of health not fully reflected by traditional risk factors such as diabetes or hypertension, including physical fitness, inflammation, and subclinical cardiovascular burden. This broader physiological signal may improve risk stratification, particularly in diverse populations where conventional risk factors alone may not fully explain differences in dementia risk. Furthermore, its inclusion may enhance model performance by capturing non-linear relationships and interactions with other cardiometabolic variables, which are more effectively leveraged by flexible machine learning approaches such as random survival forest.^32^

Another important finding is the observed variation across racial groups in predictor importance and time-to-dementia, which highlights potential heterogeneity in disease etiology and clinical presentation. These differences may reflect a complex interplay of biological, environmental, and social determinants of health, including disparities in healthcare access, socioeconomic conditions, and cumulative stress exposure.^9,10,43^ This underscores the need for race-aware and equity-informed dementia prediction models that move beyond single, uniform modeling approaches. From a methodological perspective, the strong performance of the random survival forest model highlights the advantages of flexible machine learning approaches over traditional regression methods, particularly in capturing non-linear relationships and complex interactions in heterogeneous populations. In addition to strong discrimination, its robust calibration further supports its utility for reliable risk estimation in clinical settings.

### 4.1 Strengths and Limitations

This study has several unique strengths, First, this study used of a large, longitudinal real-world cohort with over 55,000 participants and extended follow-up across two decades, enabling robust time-to-event modeling. Second, we conducted stratified analyses across four racial groups and integrated resting heart rate into models across three distinct time points, allowing evaluation of model performance and predictor importance in diverse populations. Third, the study compares random survival forest with traditional regression-based model, supported by both discrimination and calibration assessment, enhancing robustness and clinical interpretability. However, several limitations should be considered when interpreting these findings. First, NACC is not a population-representative cohort, and participants are predominantly recruited through Alzheimer’s Disease Research Centers, potentially limiting generalizability. Second, the analysis was limited to four racial groups and a maximum 5-year prediction horizon, as the sample size was insufficient to support reliable estimation of 10-year outcomes. In addition, one racial group had a comparatively smaller sample size (see Supplementary Table S5). Future studies will assess the robustness of these findings using cohorts with longer follow-up periods. Third, the dataset contained a substantial proportion of missing data; although multiple imputation was applied to reduce potential bias, residual uncertainty may still remain. Fourth, external validation in independent cohorts remains necessary before clinical implementation.

## 5 CONCLUSIONS

This study demonstrates that Random Survival Forest models using 2024 Lancet Commission risk factors and resting heart rate provide accurate dementia risk prediction across diverse racial groups. The relative importance of dementia risk factors differed substantially between racial groups, suggesting heterogeneous dementia risk profiles. Resting heart rate provided complementary prognostic information in addition to the established Lancet Commission risk factors and may support more equitable and personalized dementia risk prediction in diverse populations.

## Data Availability

The data used in this study were obtained from the National Alzheimer Coordinating Center (NACC) Uniform Data Set (UDS), Version 3.0. Data generated from the analyses performed in this study are available from the corresponding author upon reasonable request.

## Acknowledgement

The NACC database is funded by NIA/NIH Grant U24 AG072122. NACC data are contributed by the NIA-funded ADRCs: P30 AG062429 (PI James Brewer, MD, PhD), P30 AG066468 (PI Oscar Lopez, MD), P30 AG062421 (PI Bradley Hyman, MD, PhD), P30 AG066509 (PI Thomas Grabowski, MD), P30 AG066514 (PI Mary Sano, PhD), P30 AG066530 (PI Helena Chui, MD), P30 AG066507 (PI Marilyn Albert, PhD), P30 AG066444 (PI John Morris, MD), P30 AG066518 (PI Jeffrey Kaye, MD), P30 AG066512 (PI Thomas Wisniewski, MD), P30 AG066462 (PI Scott Small, MD), P30 AG072979 (PI David Wolk, MD), P30 AG072972 (PI Charles DeCarli, MD), P30 AG072976 (PI Andrew Saykin, PsyD), P30 AG072975 (PI David Bennett, MD), P30 AG072978 (PI Neil Kowall, MD), P30 AG072977 (PI Robert Vassar, PhD), P30 AG066519 (PI Frank LaFerla, PhD), P30 AG062677 (PI Ronald Petersen, MD, PhD), P30 AG079280 (PI Eric Reiman, MD), P30 AG062422 (PI Gil Rabinovici, MD), P30 AG066511 (PI Allan Levey, MD, PhD), P30 AG072946 (PI Linda Van Eldik, PhD), P30 AG062715 (PI Sanjay Asthana, MD, FRCP), P30 AG072973 (PI Russell Swerdlow, MD), P30 AG066506 (PI Todd Golde, MD, PhD), P30 AG066508 (PI Stephen Strittmatter, MD, PhD), P30 AG066515 (PI Victor Henderson, MD, MS), P30 AG072947 (PI Suzanne Craft, PhD), P30 AG072931 (PI Henry Paulson, MD, PhD), P30 AG066546 (PI Sudha Seshadri, MD), P20 AG068024 (PI Erik Roberson, MD, PhD), P20 AG068053 (PI Justin Miller, PhD), P20 AG068077 (PI Gary Rosenberg, MD), P20 AG068082 (PI Angela Jefferson, PhD), P30 AG072958 (PI Heather Whitson, MD), P30 AG072959 (PI James Leverenz, MD)

## CONFLICT OF INTEREST

None

## Notes

### Competing Interest Statement

The authors have declared no competing interest.

### Author Declarations

Health Sciences Research Ethics Board (REB) of Brock University gave ethical approval for this work (Health Sciences Research Ethics Board, REB #24072). This study used de-identified data from the NACC Uniform Data Set (UDS) version 3.0, accessed through an approved NACC data request. The NACC data were collected by participating Alzheimer Disease Research Centers (ADRCs), each of which obtained informed consent from participants and approval from its local Institutional Review Board. All data were de-identified prior to analysis.

