## Supplementary data for "Race-Specific Dementia Risk Prediction Using 2024 Lancet Commission Risk Factors and Resting Heart Rate: A Survival Analysis of 55,004 NACC Participants"

**Table S1:** Checklist of items for transparent reporting of a multivariable prediction model for individual prognosis or diagnosis (TRIPOD)

| Section/topic | Item | Development or validation | Checklist item | Page |
| --- | --- | --- | --- | --- |
| Title and abstract |  |  |  | 1 |
| Title | 1 | D;V | Identify the study as developing and/or validating a multivariable prediction model, the target population, and the outcome to be predicted | 1 |
| Abstract | 2 | D;V | Provide a summary of objectives, study design, setting, participants, sample size, predictors, outcome, statistical analysis, results, and conclusions | 2 |
| Introduction |  |  |  |  |
| Background and objectives | 3a | D;V | Explain the medical context (including whether diagnostic or prognostic) and rationale for developing and validating the multivariate prediction model, including references to existing models | 3,4 |
|  | 3b | D;V | Specify the objectives, including whether the study describes the development or the validation of the model, or both | 4 |
| Methods |  |  |  |  |
| Source of data | 4a | D;V | Describe the study design or source of data (for example, randomized trial, cohort, or registry data), separately for the development and validation datasets, if applicable | 4 |
|  | 4b | D;V | Specify the key study dates, including start of accrual; end of accrual; and if applicable, end of follow-up | 4 |
| Participants | 5a | D;V | Specify key elements of the study setting (e.g., primary care, secondary care, general population) including number and location of centres | 4 |
|  | 5b | D;V | Describe eligibility criteria for participants | 4 |
|  | 5c | D;V | Give details of treatment received, if relevant | NA |
| Outcome | 6a | D;V | Clearly define the outcome that is predicted by the prediction model, including how and when assessed | 5 |
|  | 6b | D;V | Report any actions to blind assessment of the outcome to be predicted | NA |
| Predictors | 7a | D;V | Clearly all predictors used in developing the multivariate prediction model, including how and when they assessed | 5,6 |
|  | 7b | D;V | report any actions to blind assessment for predictors for the outcome and other predictors | NA |
| Sample size | 8 | D;V | Explain how the study size was arrived at | 4 |
| Missing data | 9 | D;V | Describe how missing data were handled (for example, complete-case analysis, single imputation, multiple imputation) with details of any imputation method | 6, FS1 |
| Statistical analysis methods | 10a | D | Describe how predictors were handled in the analyses | 6 |
|  | 10b | D | Specify type of model, all model-building procedures (including any predictor selection), and method for internal validation | 6-7 |
|  | 10c | V | For validation, describe how the predictions were calculated | 6-7 |
|  | 10d | D;V | Specify all measures used to assess model performance and, if relevant, to compare multiple models | 6-7 |
|  | 10e | V | Describe any model updating (for example, recalibration) arising from the validation, if done | NA |
| Risk groups | 11 | D;V | Provide details on how risk groups were created, if done | NA |
| Development v validation | 12 | V | For validation, identify any differences from the development data in setting, eligibility criteria, outcome, and predictors | 9-10 |
| Results |  |  |  |  |
| Participants | 13a | D;V | Describe the flow of participants through the study, including the number of participants with and without the outcome and, if applicable, a summary of the follow-up time. A diagram may be helpful | Table 1&2 |
|  | 13b | D;V | Describe the characteristics of the participants (basic demographics, clinical features, available predictors), including the number of participants with missing data for predictors and outcome | 7, Table 1&2 |
|  | 13c | V | For validation, show a comparison with the development data of the distribution of important variables (demographics, predictors, and outcome) | 8 |
| Model development | 14a | D | Specify the number of participants and outcome events in each analysis | Table 1&2 |
|  | 14b | D | If done, report the unadjusted association between each candidate predictor and outcome d | Figure 1 |
| Model specification | 15a | D | Present the full prediction model to allow predictions for individuals (that is, all regression coefficients, and model intercept, or baseline survival at a given time point) | NA |
|  | 15b | D | Explain how to use the prediction model | 9 |
| Model performance | 16 | D;V | Report performance measure (with CIs) for the prediction model | 8 |
| Model updating | 17 | V | If done, report the results from any model updating (that is, model specification, model performance) | Figure 1-4 |
| Discussion |  |  |  |  |
| Limitations | 18 | D;V | Discuss any limitations of the study (such as nonrepresentative sample, few events per predictor, missing data) | 11 |
| Interpretation | 19a | V | For validation, discuss the results with reference to performance in the development data, and any other validation data | 10,11 |
|  | 19b | D;V | Given an overall interpretation of the results, considering objectives, limitations, results from similar studies, and other relevant evidence | 10,11 |
| Implications | 20 | D;V | Discuss the potential clinical use of the model and implications for future research | 11 |
| Other information |  |  |  |  |
| Supplementary information | 21 | D;V | Provide information about the availability of supplementary resources, such as study  protocol, Web calculator, and data sets | NA |
| Funding | 22 | D;V | Give the source of funding and the role of the funders for the present study | 26 |

*Items relevant only to the development of a prediction model are denoted by D, items relating solely to a validation of a prediction model are denoted by V, and items relating to both are denoted D;V.

**Table S2. NACC variables used for dementia prediction and corresponding variable definitions**

| Variable Name | Descriptor |
| --- | --- |
| RACE | Race |
| HRATE | Resting heart rate (pulse) |
| SEX | Subject sex |
| EDUC | Years of education |
| NACCBMI | Body Mass Index (BMI) |
| NACCAGE | Subject age at visit |
| DEMENTED | Subjects who met the criteria for all-cause dementia |
| BRNINJ | Traumatic brain injury |
| HYPERT | Hypertension present |
| NACCFDYS | Days from initial visit to each follow-up visit |
| MOCAHEAR | Subject was unable to complete one or more sections due to hearing |
| HYPCHOL | Hypercholesterolemia present |
| NACCADEP | Reported current use of an antidepressant |
| DIABET | Diabetes present at visit |

**Table S3: Integrated Brier Scores of dementia prediction models across racial groups**

| Random Survival Forest | | | | |
| --- | --- | --- | --- | --- |
| Survival Threshold | White | Black | Asian | American  Indian |
| One-year | 0.02 | 0.02 | 0.02 | 0.02 |
| Three-year | 0.12 | 0.09 | 0.11 | 0.12 |
| Five -year | 0.13 | 0.12 | 0.14 | 0.13 |
| Cox Proportional Hazard | | | | |
| One-year | 0.01 | 0.01 | 0.01 | 0.02 |
| Three-year | 0.11 | 0.08 | 0.09 | 0.11 |
| Five -year | 0.13 | 0.12 | 0.15 | 0.14 |

**Table S4: Integrated Brier Scores of dementia prediction models developed without resting heart rate**

| Random Survival Forest | | | | |
| --- | --- | --- | --- | --- |
| Survival Threshold | White | Black | Asian | American  Indian |
| One-year | 0.02 | 0.02 | 0.02 | 0.02 |
| Three-year | 0.12 | 0.09 | 0.11 | 0.12 |
| Five -year | 0.13 | 0.12 | 0.15 | 0.12 |
| Cox Proportional Hazard | | | | |
| One-year | 0.01 | 0.01 | 0.01 | 0.02 |
| Three-year | 0.11 | 0.08 | 0.09 | 0.11 |
| Five -year | 0.13 | 0.12 | 0.15 | 0.14 |

**Table S5. Distribution of dementia and non-dementia participants across racial groups and prediction horizons (1-, 3-, 5-, and 10-Year).**

|  | 1 – year  11,520/7723 | 3- years  7,594/5990 | 5 -years  10,269/7268 | 10- years  3517/1123 |
| --- | --- | --- | --- | --- |
| White | | | | |
| No Dementia | 8,065 (70%) | 5,835 (77%) | 8,173 (80%) | 2,886 (82%) |
| Dementia | 6,471 (84%) | 5,189 (87%) | 6,419 (88%) | 967 (86%) |
| Black | | | | |
| No Dementia | 2,780 (24%) | 1,420 (19%) | 1,706 (17%) | 524 (15%) |
| Dementia | 971 (13%) | 607 (10%) | 652 (9.0%) | 127 (11%) |
| Asia | | | | |
| No Dementia | 460 (4.0%) | 240 (3.2%) | 295 (2.9%) | 92 (2.6%) |
| Dementia | 190 (2.5%) | 127 (2.1%) | 155 (2.1%) | 27 (2.4%) |
| American Indian | | | | |
| No Dementia | 203 (1.8%) | 93 (1.2%) | 83 (0.8%) | 14 (0.4%) |
| Dementia | 74 (1.0%) | 56 (0.9%) | 37 (0.5%) | 1 (<0.1%) |
| Native Hawaiian | | | | |
| No Dementia | 12 (0.1%) | 6 (<0.1%) | 12 (0.1%) | 1 (<0.1%) |
| Dementia | 17 (0.2%) | 11 (0.2%) | 5 (<0.1%) | 1 (<0.1%) |

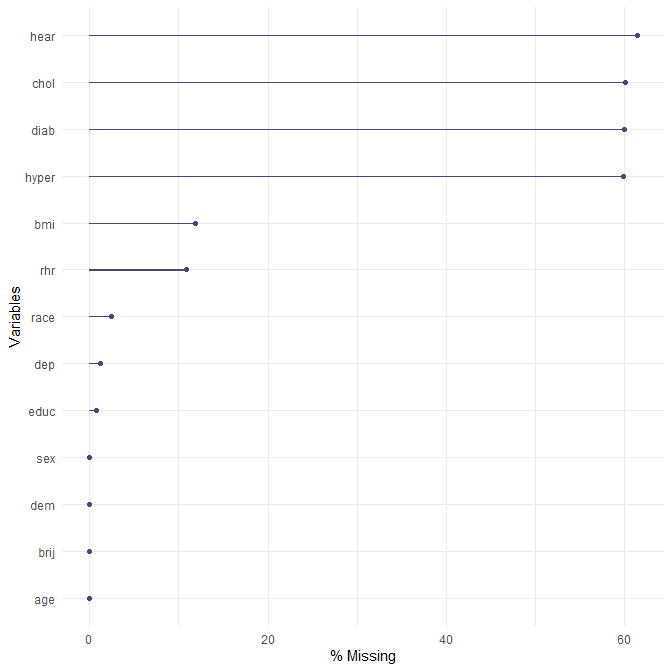

**Figure S1. Distribution of missing data across study variables.** The percentage of missing observations is shown for each predictor included in the dementia prediction models. Hypercholesterolemia, diabetes, hypertension, and hearing loss exhibited the highest levels of missingness, supporting the use of multiple imputation to reduce potential bias and maximize data utilization. Variables shown on the y-axis include age, sex, education, body mass index (BMI), resting heart rate (RHR), hypercholesterolemia, hypertension, depression, diabetes, traumatic brain injury, and hearing loss.

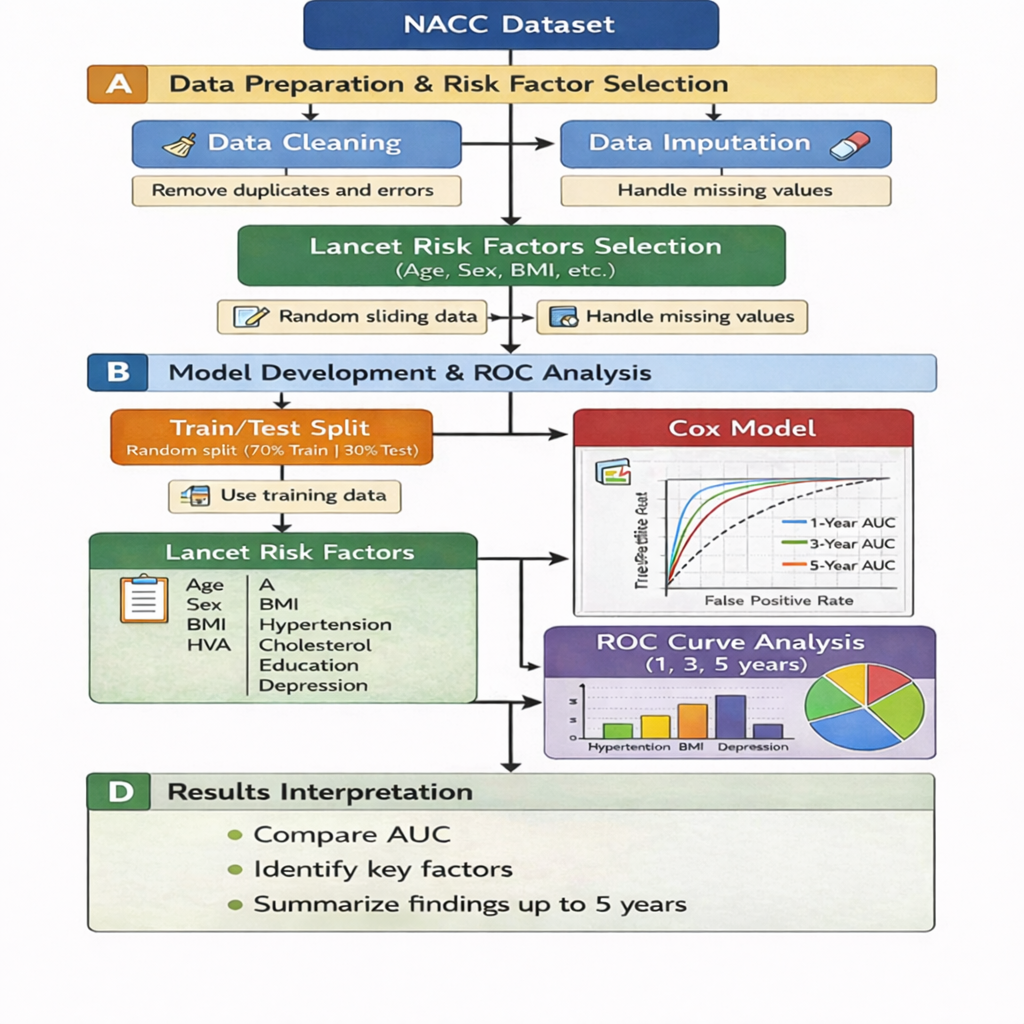

**Figure S2. Workflow of data preprocessing, model development, and performance evaluation.** Longitudinal NACC records were transformed into participant-level survival datasets, followed by data cleaning, multiple imputation, predictor selection, model training, and evaluation using Cox proportional hazards and Random Survival Forest models. Predictive performance was assessed using time-dependent AUC and Integrated Brier Score.

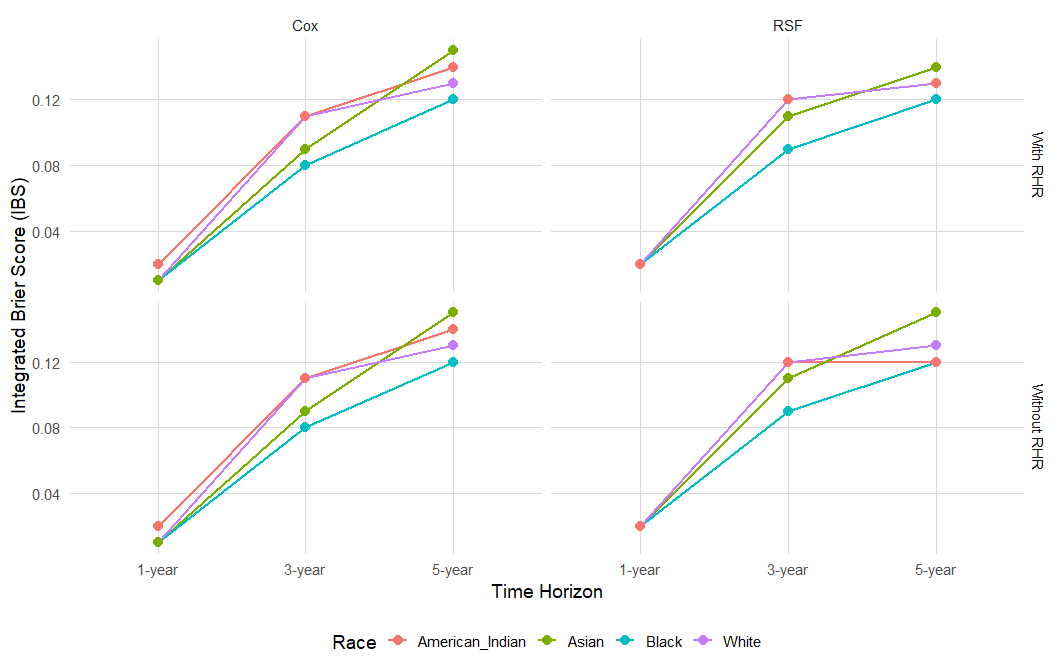

**Figure S3. Calibration performance assessed using Integrated Brier Scores across racial groups.** Integrated Brier Scores for Cox proportional hazards and Random Survival Forest (RSF) models are shown with and without inclusion of resting heart rate (RHR). Lower IBS values indicate better overall predictive performance. Random Survival Forest models maintained consistently low IBS values across racial groups and prediction horizons.

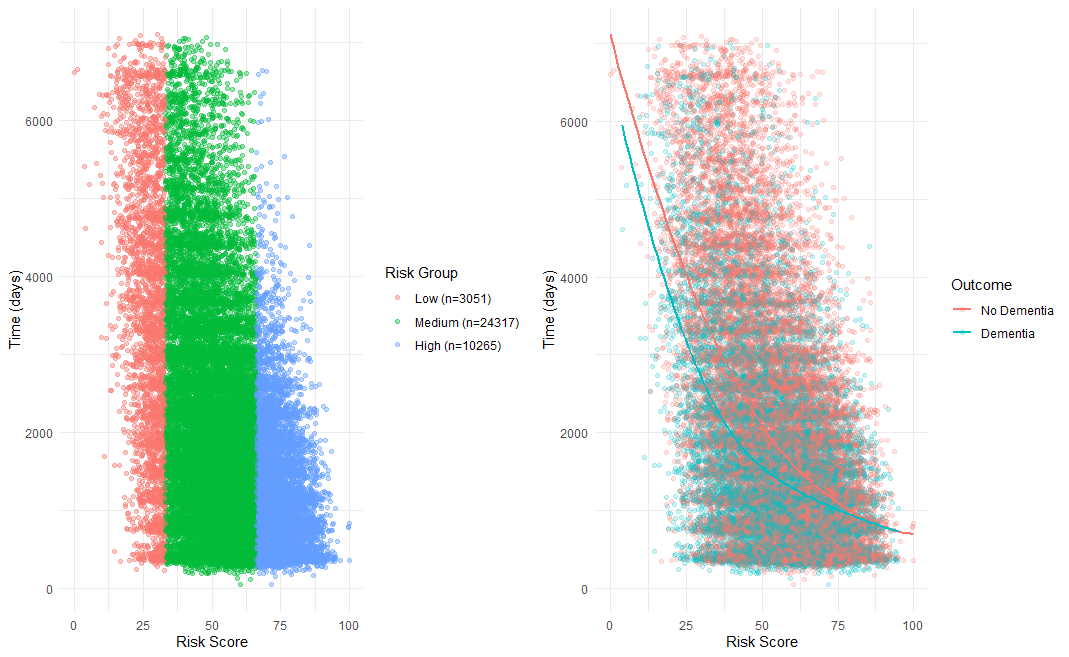

**Figure S4. Dementia risk stratification using model-derived risk scores.** Left panel: distribution of participants classified into low-, medium-, and high-risk groups according to predicted dementia risk. Right panel: relationship between predicted risk score and time to dementia onset. Individuals with higher risk scores tended to develop dementia earlier, whereas lower risk scores were associated with prolonged dementia-free survival, supporting the clinical utility of model-based risk stratification.

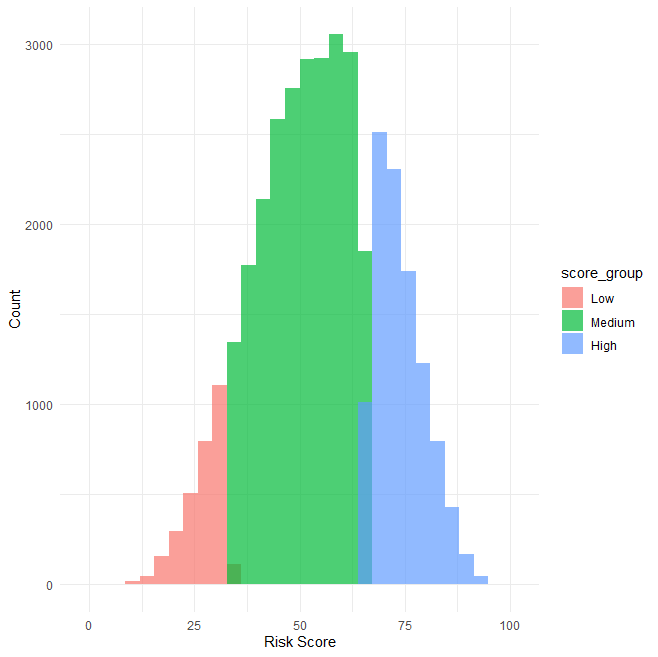

**Figure S5. Distribution of model-derived dementia risk scores.** Predicted risk scores were approximately normally distributed and were subsequently categorized into low-, medium-, and high-risk groups for risk stratification analyses.
